# Editors and research articles: a retrospective study of self-publication in biomedical research

**DOI:** 10.64898/2026.08.17.26360597

**Authors:** Cédric Lemarchand, Florian Naudet, Marc-Antoine Pencolé, Louën Ropers, Alexandre Scanff, Ioana Alina Cristea, Clara Locher

## Abstract

**Objective:** In 2021, a large-scale survey highlighted that in a subset of biomedical journals, a few authors –often serving on the editorial board– published disproportionately and experienced shorter acceptance times. Our study aims to specifically quantify editors’ *research articles* within the journals in which they operate.

**Methods:** We selected journals indexed in Open Editors, a dataset that collects publicly available information on journal editorial boards through web scraping. Journals not indexed in PubMed, mega-journals, and those with very low publication volume were excluded. For the remaining journals, we linked the 2022 editorial boards from Open Editors to authors of *research articles* (i.e., original articles, case reports, and reviews) published between 2020 and 2023. For each journal, we then computed indicators describing publication patterns: the percentage of *research articles* (i) by the most prolific editor, (ii) with at least one editor, and (iii) by the most prolific author, as well as publication lags for each article.

**Results:** Across the 1,623 journals studied, the median and 95^th^ percentile of *research* articles are 1.78% and 6.7% for those co-authored with the most prolific editor, 12.1% and 41.1% for those with at least one editor, and 2.5% and 7.9% for those with the most prolific author. An editor was among the most prolific author(s) in 45.0% of the journals. For authors, the median and 5^th^ percentile publications lags are 99 and 35 days; for editors, it is 95 and 33 days; and for editors-in-chief, it amounts to only 84 and 12 days. An in-depth examination of journals where the most prolific editor co-authored more than 6.7% (95^th^ percentile) found a median impact factor of 3, and a median h-index of 42 for their most prolific editor(s).

**Conclusion:** In 5% of cases, an editor contributes to approximately >7% of the articles published in their own journal. In nearly half of the journals, the most prolific author is an editor. These results need to be complemented by a qualitative approach to examine whether *research articles* authored by editors appropriately address potential conflicts of interest, as required by COPE recommendations, and to better understand the motivations underlying this practice.

## Introduction

Scientific journal editors play the crucial role of gatekeepers for the publication of scientific content. Consequently, flaws in editorial practices may directly affect the trustworthiness of the published evidence.

With the rise of predatory journals – a well-known and well-researched phenomenon (1,2) – and the growing recognition of fake manuscripts submission (3), ensuring the integrity of the publication process is more crucial than ever to preserve the validity of scientific literature. More broadly, given the prevalence of research misconduct, which finds some of its roots in academia (4), research integrity has emerged as a legitimate topic on its own. Accordingly, emerging tools to identify and quantify questionable research practices are being progressively developed (5,6) and problematic patterns of published articles, such as tortured phrases, have recently come under scrutiny (7).

While most efforts have focused on journals flagged with major integrity issues, less is known about merely questionable editorial practices that may also occur in otherwise “legitimate” journals. Among these practices, we highlight self-publication, that could be defined as the publication of a research article by a member of the editorial board, in their own journal. The Committee on Publication Ethics (COPE) stated in one of its cases (8) that self-publication might be legitimate in the appropriate circumstances, “particularly in those circumstances where the choice of journals is limited”. However, although COPE acknowledges that potential bias cannot be completely removed even if the editor is not involved in the peer-review process, COPE recommends that such cases of self-publication should be transparently disclosed.

To better characterize the phenomenon of self-publication, a survey (9) explored the relevance of two indicators to identify journals with self-promotional tendencies: the percentage of paper by the most prolific author (PPMP) and the Gini coefficient (10), traditionally used in economics as a measure of statistical dispersion to represent income, wealth or consumption inequalities. Articles authored by the most prolific author(s) were more likely to be accepted within three weeks (10.2% against 1.9%). Further analysis highlighted an overrepresentation of editorial board members amongst the most prolific authors. This gap may suggest differences of treatment of manuscripts submitted by most prolific authors and remaining authors, and/or between authors and editors. Similar tendencies were backed by another study (11) on Elsevier journals. In addition, a systematic review (12) of studies exploring editors publishing in their own(s) journal(s) suggested high levels of self-publication compromised by methodological flaws.

Taken together, these findings raise concerns that recurring self-publication may create situations of favoritism, whereby self-publishing authors benefit from shorter acceptance times. Such accelerated publication rates may contribute to inflated publication metrics at both the author and journal levels. More importantly, it may suggest that peer-review procedures are, at least in some cases, less rigorous than expected.

In this context, our objective is to provide a comprehensive quantification of self-publication occurrences within the biomedical literature, including the exploration of indicators that may detect them accurately.

## Material & Methods

The research protocol was prospectively registered on the Open Science Framework (OSF) platform (13) prior to any data collection on March 21^st^, 2025. Any deviation from the registered protocol was transparently reported.

### Search strategy

To provide a comprehensive quantification of self-publication occurrences within the biomedical literature, we had to chain two separate data sources: Open Editors (14) and PubMed.

Open Editors collects data on editorial boards through R-written web scraping. The 2022 dataset, the most recent available at the time of our study, compiled from scraping rounds, comprises 594,580 editors, 7,352 journals, and 26 publishers. The extracted information includes editor’s name, affiliation, journal (including International Standard Serial Number (ISSN)), publisher and role. A subset of Open Editors’ dataset restricted to the selection criteria described below was used for data extraction. Then, publications were harvested using a dedicated R-written script. We used the easyPubMed 3.1.3 (15), a R-developed package designed to facilitate the extraction and aggregation of data from the E-utilities API. The script extracts all articles published in the journals covered by Open Editors’ dataset and available on PubMed, within the time frame specified in our inclusion criteria (2020 to 2023). For each journal, the query used the ISSN and the time frame: *“ISSN”[ta] AND 2020:2023[dp]*, with additional refinement explicated below.

### Selection criteria

#### Journal selection

We considered journals (a) covered by Open Editor’s dataset, (b) with an entered ISSN either for their physical or digital edition and (c) indexed in PubMed. In those journals, articles data were extracted from January 1, 2020, to December 31, 2023. This time frame represents a four-years interval centered around the web scraping’s date, between January and February 2022. The selection of a four-years interval rests on two hypothesis : it is quite likely that actual editors in 2022 were already occupying the same position in the past two years, and kept their seat in the following two years; were it not the case, it would still be reasonable to expect a certain degree of interpersonal relationship between the future/former editor and the board members up to two years before or after, relationship that would expose to the possibility of a clientelist publication bias.

To ensure a sufficient sample size for each journal included in the analysis, only journals with at least 50 research articles during the study period were considered. On the opposite end of the spectrum, mega-journals were excluded due to higher risks of author homonymy, longer extraction times, and a publication volume that highly differs from those of smaller journals and could significantly alter our analysis parameters. Ioannidis et al. define them using specific criteria, one of them being a quantity of more than 2000 articles per year (16).

#### Article selection

We defined research articles as publications that provide original knowledge (i.e., original articles, case reports and reviews), and do not consist in journalistic activities. Since PubMed does not have a specific Publication Type that allows for the identification of research articles (i.e., original articles, case reports and reviews), research articles were defined in a negative way. This approach, consisting in including articles with ‘publication type’ corresponding to ‘journal article’ and excluding articles without abstracts, was already used in previous research (10,16,17), thus wasn’t modified for this study. Additionally, articles were excluded if falling under the following tags for ‘publication type’, as defined in Scanff et al. protocol: ‘Comment’, ‘Letter’, ‘Editorial’, ‘Published Erratum’, ‘News’, ‘Introductory Journal Article’, ‘Biography’, ‘Portrait’, ‘Congress’, ‘Interview’, ‘Retraction of Publication’, ‘Personal Narrative’, ‘Retracted Publication’’, ‘Patient Education Handout’, ‘Lecture’, ‘Autobiography’, ‘Clinical Conference’, ‘Classical Article’, ‘Address’, ‘Legal Case’, ‘Expression of Concern’, ‘Festschrift’, ‘Overall’, ‘Bibliography’, ‘Corrected and Republished Article’, ‘Interactive Tutorial’, ‘Duplicate Publication’, ‘Directory’, ‘Newspaper Article’, ‘Periodical Index’, ‘Dictionary’.

### Disambiguation of authors’ and editors’ names

For each article, the authors’ list was matched with the list of editors for the journal, as compiled by Open Editors in 2022. This represents a necessary step since the authors’ names (extracted from PubMed) and the editors’ names (extracted from Open Editors) are structured differently: while Open Editors features a single column for author’s name, easyPubMed permits extraction of the first and last names separately. To ensure the reliability of the obtained data without a specific identifier for each editor, we proceeded to disambiguation in multiple steps. Before any further processing, the dataset was harmonized by converting all text to lowercase and normalizing all special characters.

Additionally, we decided to exclude multiple occurrences of a single editor’s name that were impossible to discriminate (i.e. “J. Smith” and another “J. Smith”). We thus prioritized specificity over sensibility.

### Filter validation

Preliminary tests were run to describe the performances of the filters: identification of research article in PubMed, disambiguation between author and editor names, and identification of editors-in-chief in Open Editors. For each filter, the reference standard was a manual extraction performed by either one (CLe) or two reviewers (FN or CLo).

A first sample of 100 randomly selected articles was taken from a dataset with all available articles on PubMed from the 1,623 selected journals, including research articles. Two independent reviewers (CLe and FN/CLo) identified the research articles in the subset, with a third reviewer (FN/CLo) to decide between two contradictory answers in potentially ambiguous cases. The results between the filter and the manual selection were analyzed in a confusion matrix (with actual condition being the results of the manual selection and predicted condition being the results of the filter). The editors-in-chief and disambiguation filters were tested on a sample of editorial roles within a sample of 25 journals, with a single reviewer’s extraction (CLe). To set up the disambiguation filter, we ensured that we had sufficient data by selecting cases where at least PubMed’s last name was found in Open Editors’ name field.

For all filters, explored outcomes for the preliminary tests are sensibility (probability that an actually *positive* case was correctly identified by the filter), specificity (probability that an actually *negative* case was correctly identified by the filter), positive predictive value [PPV] and negative predictive value [NPV], with specificity as our main outcome. Our target specificity was an arbitrary 80%, to limit the risks of false positives. Below this threshold, it was planned to adjust the filters and repeat the validation.

### Indicators

Our primary outcome was the Percentage of Papers by the Most Prolific Editor (PPMPE). It corresponds to the proportion of papers by the most prolific editor relative to total papers (**Table 1A**). To complete the information provided by the PPMPE, two complementary indicators were used: the Percentage of Papers with at least One Editor (PPOE), and the Percentage of Papers by the Most Prolific author (PPMP), that follow the same calculation model (**Table 1A**).

**Table 1:**
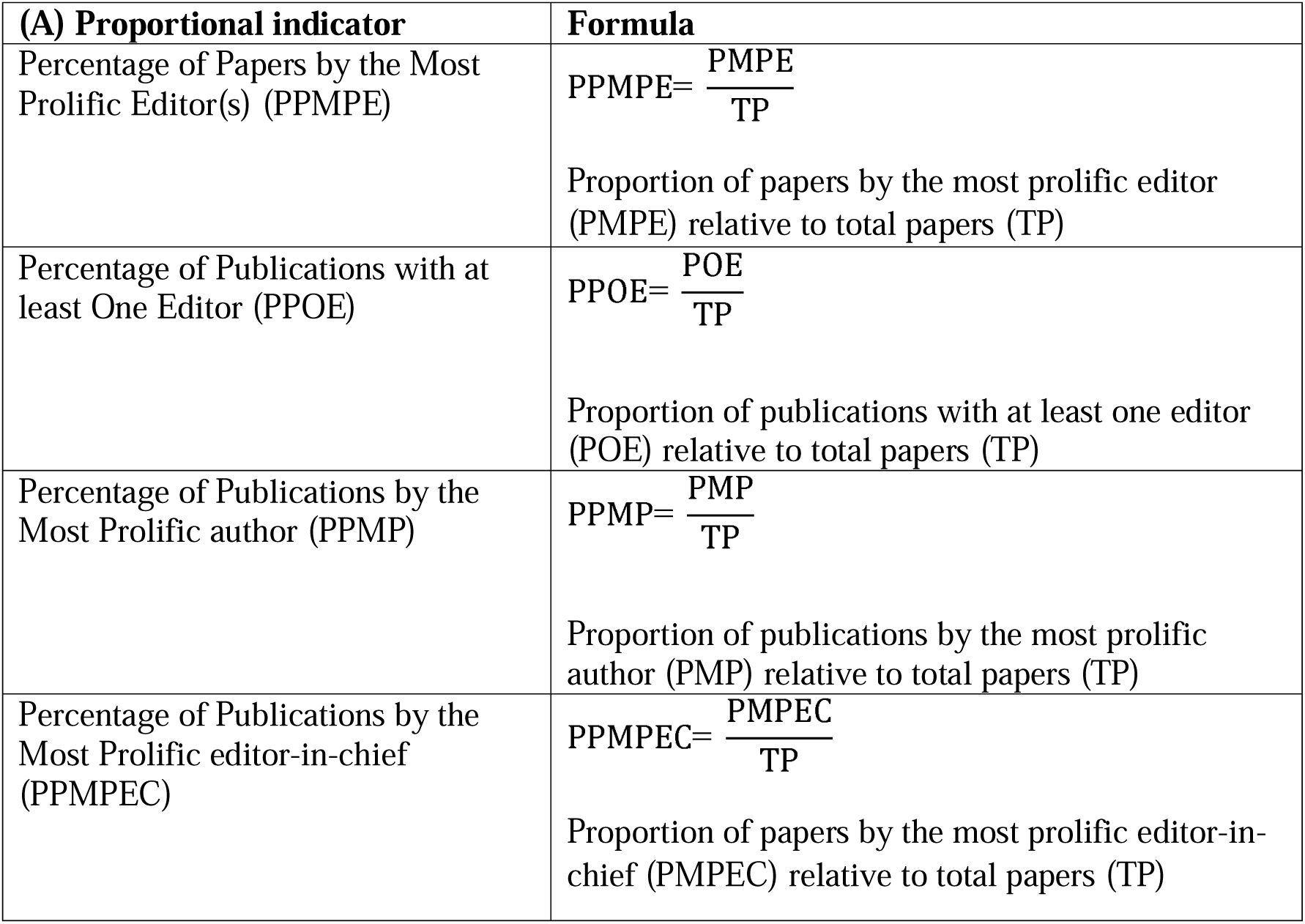
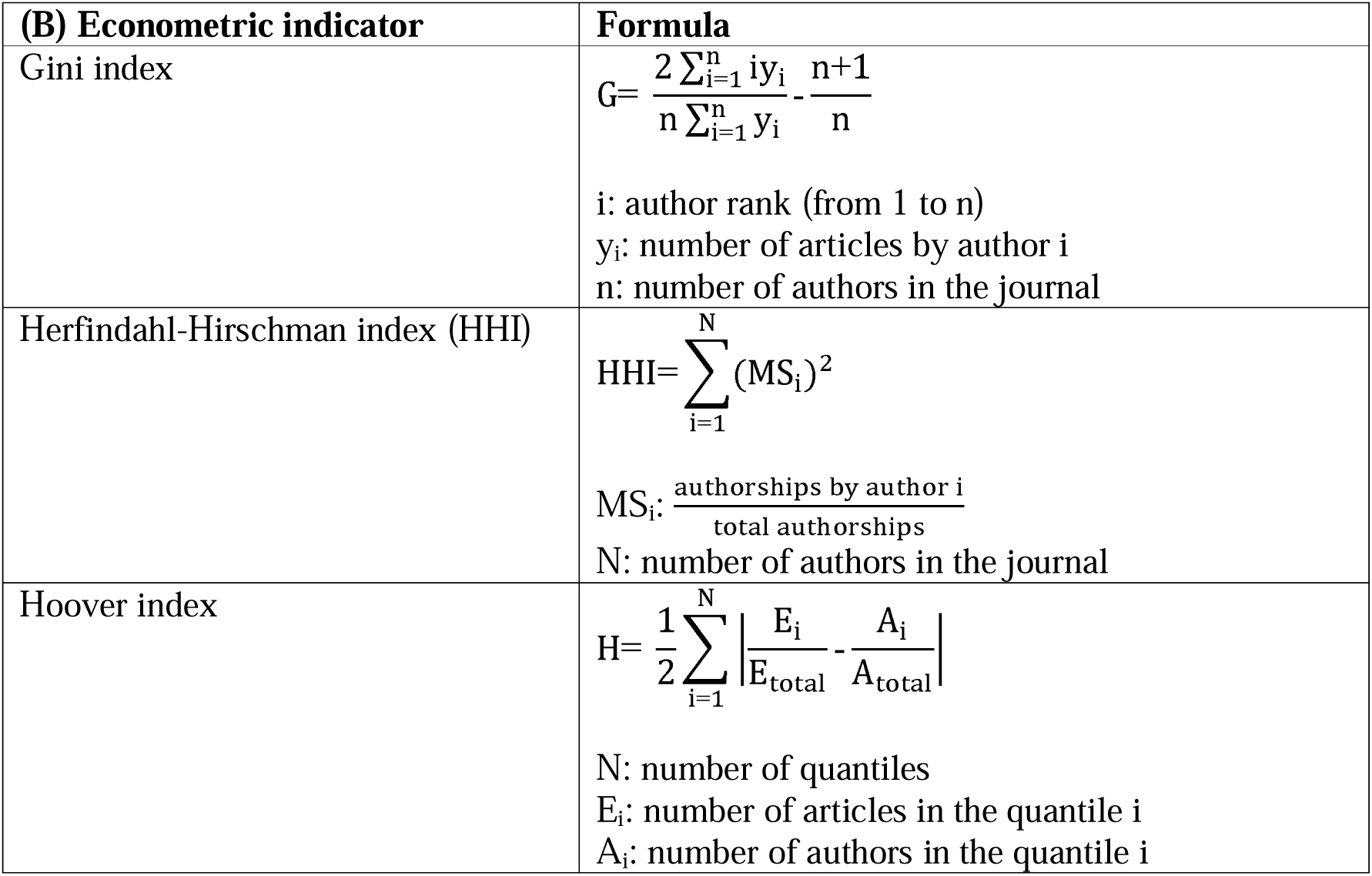
Proportional and econometric indicators used to measure self-publication, and their respective formulas.

To assess differences in the rhythm of publication between journals, or articles, we used the publication lag (PL). This publication lag is defined by the number of days between the submission date, and the publication date.

To complete this analysis, and provide information not only on individual authors and editors, but groups of authors as well, we are also bringing econometric indicators (**Table 1B**). These econometric indicators are usually employed to address the disparities between the resources owned by a country’s inhabitants. In our study, the owned resource was the authorship.

The Gini index (**Supplementary fig. 1**) is often used in econometrics to describe resource distribution inequalities. This measure of statistical dispersion has a range between 0 (perfect equality) and 1 (maximal inequality). It may be used in several bibliometric studies to explore authorship inequalities (17,9,18). Similarly, the less common Hoover index represents the percentage of the total population’s resources that should be redistributed in order to achieve perfect equality. Graphically, these two indicators can be represented with a Lorenz curve, a graphical representation of the distribution of a resource within a population.

On the other hand, the Herfindahl-Hirschman index (**Supplementary fig. 2**) measures the market share within an industry. In the context of publication, it may be used to measure the authorship share within a journal. Its range also spans between 0 (equal repartition of the share between firms) and 1 (monopolistic situation).

### Analysis

A descriptive analysis of all parameters was performed, following their distribution (mean and standard deviation for normal distributions, median and interquartile range for other distributions). Additionally, for each parameter, the 5^th^ and 95^th^ percentiles were indicated. We provided further insight on the outliers (h-index of the most prolific editor(s) and impact factor of the journal), defined as being above the upper 95^th^ percentile of the PPMPE. At journal level, publication lag was computed between articles with at least one editor-in-chief, articles with at least one editor, and articles without any of the latter. Furthermore, we modelled the publication lag using a linear mixed model, treating the journal and the journal type as random effects, and editorial function as fixed effect.

We used the R 4.4.2 and Python 3.11.3 programming languages, with the RStudio and Visual Studio Code IDEs. The code and the versions of each used package are available on the OSF.

#### Changes to initial protocol

A sample of 100 journals was initially planned for the analysis of PPMPE outliers. However, with the final journal sample consisting of 1,623 journals, sampling was not needed and all of the 80 outliers were analyzed.

Editors-in-chief and disambiguation filters were tested on a sample of 25 journals, as each journal featured several items and this sample size was sufficient to get more than 100 items for each analysis. Contrary to the initial protocol, only a single reviewer (CLe) performed the extraction.

We did not analyze several outcomes originally defined in our protocol, including correction lags, retraction lags, and journal specialties, because missing data compromised the quality of the information to an extent that conventional methods could not adequately address.

## Results

### Journal selection and description

Open Editors features the editorial board composition of 7,352 journals. Of these, 1,733 journals without an ISSN and 2,416 journals not indexed in PubMed were excluded. Among the remaining 3,203 journals, 66 exceeded our upper publication threshold, and 1,514 did not meet the minimum publication threshold (50 research articles or more in 4 years, no more than 8,000 research articles in 4 years or 2,000 research articles in a year). The final dataset comprised 1,623 journals (**Supplementary fig. 3**).

Between 2020 and 2023, these journals published a total of 1,017,901 research articles with a median of 353 research articles per journal (IQR 188 to 700). Overall, 150,302 (14.8%) were co-authored by at least one editor, with a median of 42 per journal (IQR 16 to 102) (**Table 2**).

**Table 2:**
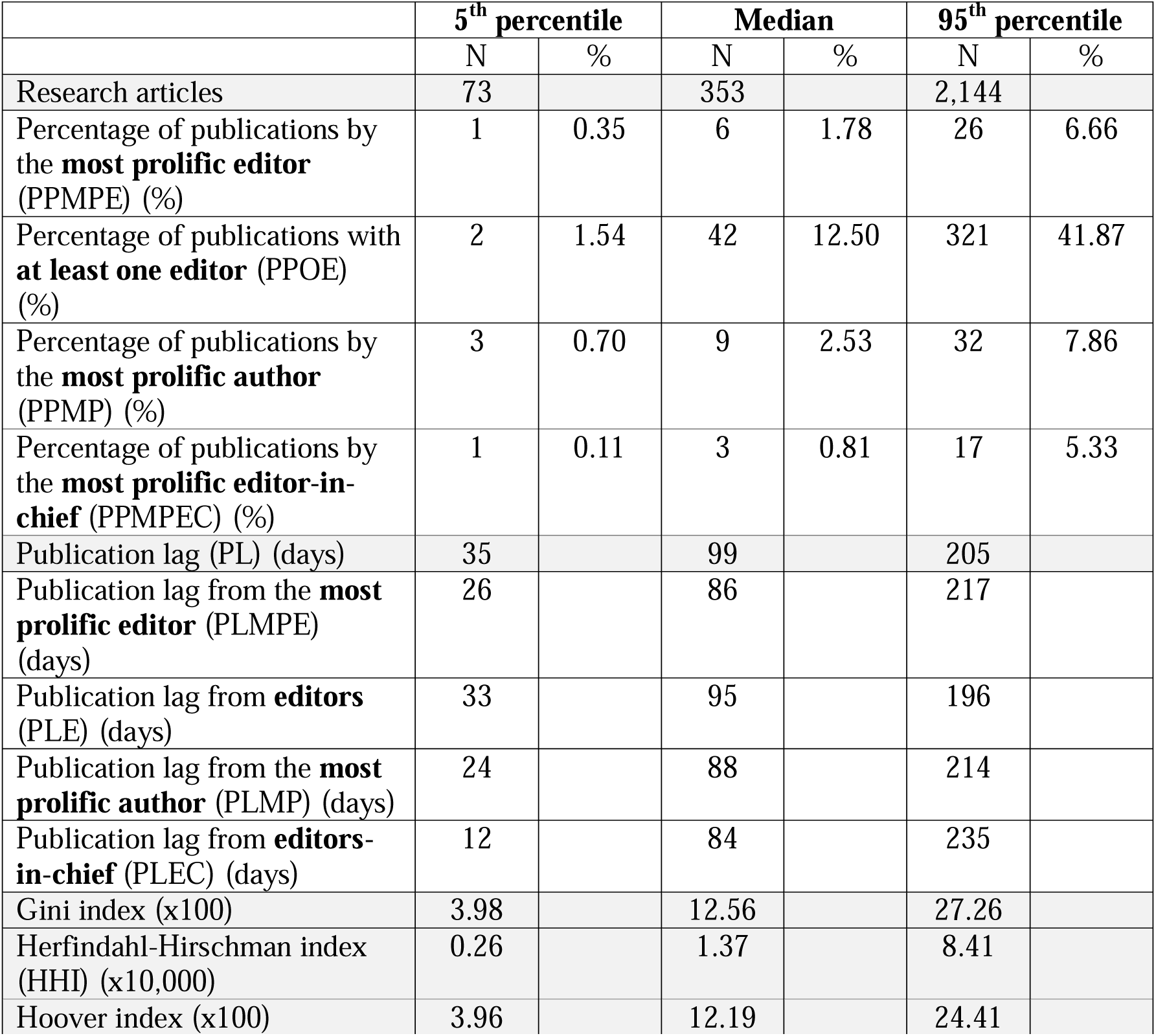
Median, 5^th^ and 95^th^ percentile of the indicators; numeric and proportional values are independent.

Distribution of authorship and editor publication indicators

## Authorships

Among the 1,623 journals, 1,583 (98%) had at least one research article co-authored by at least one editor, and 1,064 (66%) had at least one research article co-authored by an editor-in-chief. **Figure 1** summarizes the distribution of the four authorship indicators according to journal publication volume.

**Figure 1:**
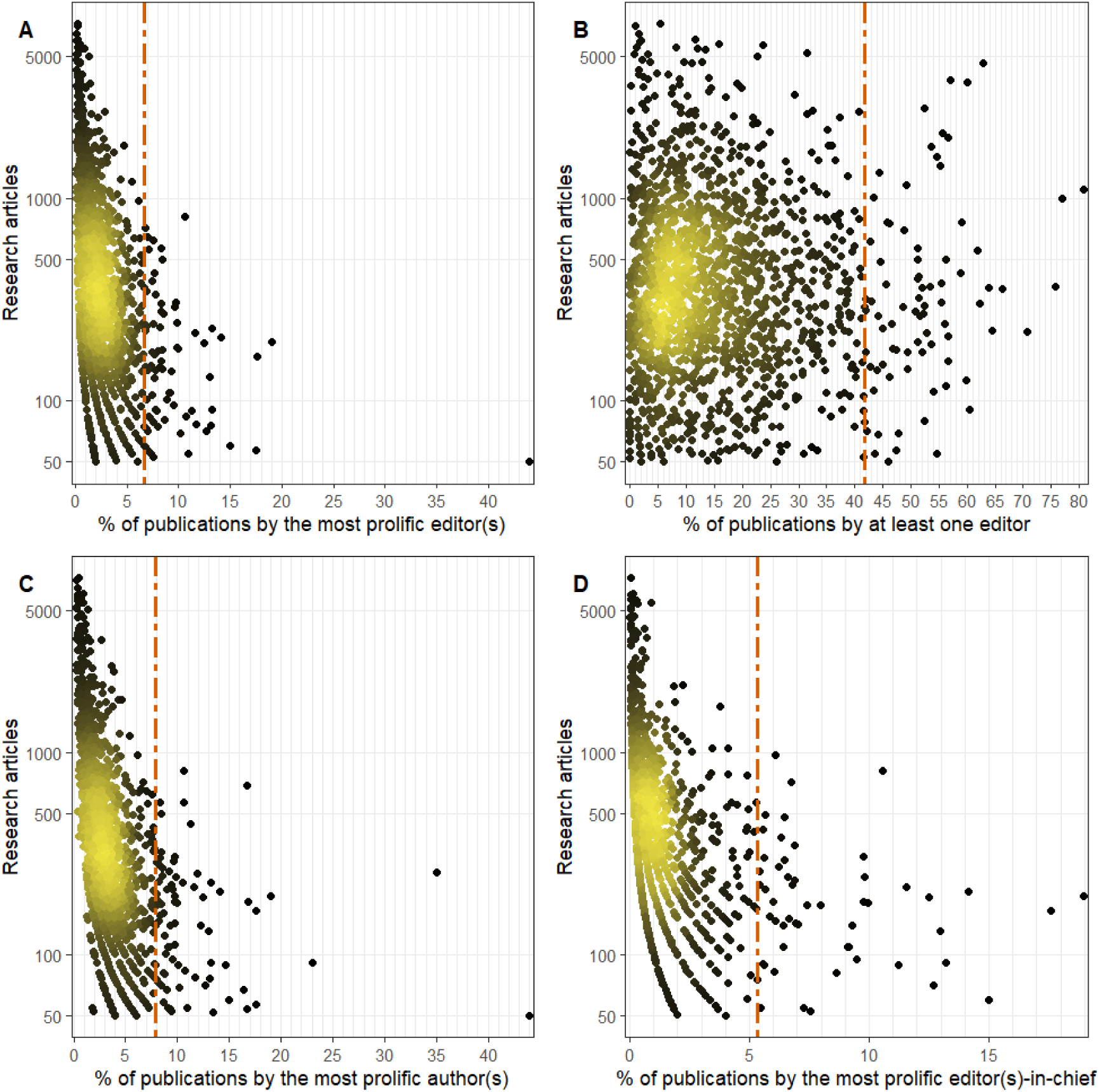
Percentage and 95^th^ percentiles of publications by the most prolific author(s), the most prolific editor(s), at least one author and the most prolific editor(s)-in-chief to total research articles between 2020 and 2023.

### Most prolific editor(s) (PPMPE)

**Fig. 1A** presents the percentage of research papers published by the most prolific editor (PPMPE) for the 1,583 journals with at least one editor identified. The median PPMPE was 1.8% (IQR 1.0% to 3.1%), with a 95^th^ percentile of 6.7%. In 357 (22.6%) journals, more than one most prolific editor was identified. The most prolific editor(s) published a median of 6 research articles (IQR 4 to 11), reaching 26 articles at the 95^th^ percentile.

### At least one editor (PPOE)

**Fig. 1B** presents the percentage of research papers co-authored by at least one editor (PPOE). Considering all editors, the median and 95^th^ percentile of research articles with at least one editor were 12.5% (IQR 6.0% to 22.2%) and 41.9%.

### Most prolific author(s) (PPMP)

**Fig. 1C** presents the percentage of research papers published by the most prolific author (PPMP). Between 2020 and 2023, the median PPMP was 2.5% (IQR 1.5% to 4.1%), with a 95^th^ percentile of 7.9%. The most prolific author(s) published a median of 9 research articles (IQR 5 to 14), reaching 32 research articles at the 95^th^ percentile. In 475 (29.3%) journals, more than one most prolific author was identified. Among the 1,623 journals, an editor was among the most prolific author(s) in 730 (45.0%) journals.

### Most prolific editor(s)-in-chief (PPMPEC)

**Fig. 1D** presents the percentage of research papers co-authored by the most prolific editor-in-chief (PPMPEC) for the 1,064 (65.6%) journals where an editor-in-chief was identified. The median PPMPEC was 0.8% (IQR 0.4% to 1.8%), with a 95^th^ percentile of 5.3%. The most prolific editor-in-chief published a median of 3 research articles (IQR 2 to 6), reaching 17 articles at the 95^th^ percentile. In 90 (8.5%) journals, more than one most prolific editor-in-chief was identified. An editor-in-chief was among the most prolific editor(s) in 313 (19.3%) of the journals, and the most prolific author(s) in 142 (8.7%) journals.

## PPMPE outliers

An in-depth examination of the 80 journals where the most prolific editor co-authored more than 6.7% (95th percentile) found a median impact factor of 3, and a median h-index of 42 for their most prolific editor(s). In this sample, 29 most prolific editors (36.3%) were editors-in-chief.

## Publication lags

**Figure 2** summarizes publication lags across journals according to authorship status. Across all authors, the median publication lag was 99 days (IQR 72 to 129), with a 5th percentile of 35 days. The most prolific editor(s) had a median publication lag of 86 days (IQR 57 to 122), with a 5^th^ percentile of 27 days (**Fig. 2A**). Among editors, the median publication lag was 95 days (IQR 69 to 126), with a 5th percentile of 33 days (**Fig. 2B**). The most prolific author(s) had a median publication lag of 88 days (IQR 57 to 122), with a 5th percentile of 24 days (**Fig. 2C**). Among editors-in-chief, the median publication lag was 84 days (IQR 51 to 126), with a 5th percentile of 12 days (**Fig. 2D**).

**Figure 2:**
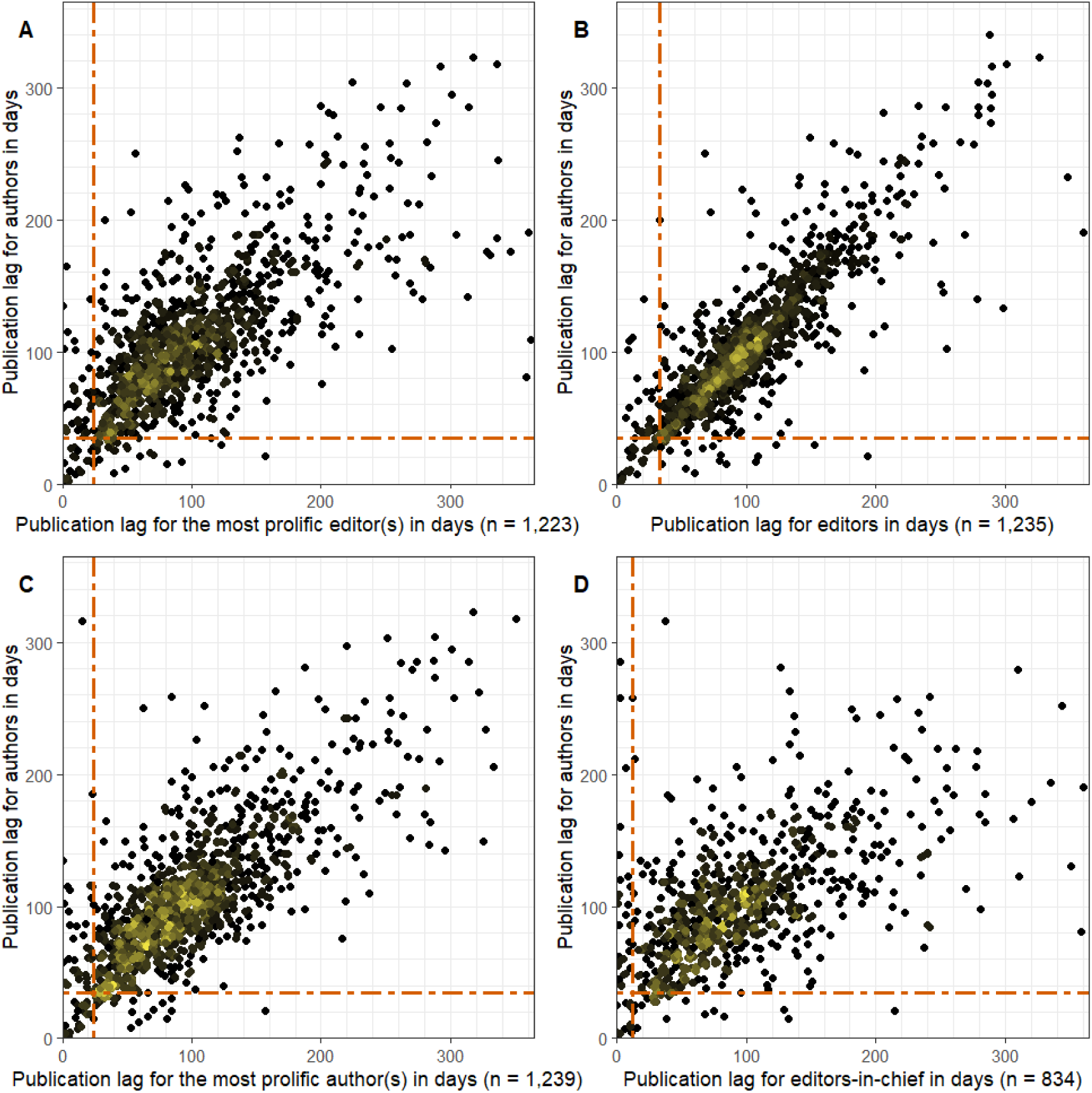
Publication lag and 5^th^ percentiles for all editors, editors-in-chief, the most prolific author(s) and the most prolific editor(s) related to publication lag for all authors between 2020 and 2023.

At article-level, using a linear mixed model separating editors from editors-in-chief, we found that an editor, on average, takes one additional day to publish compared to a non-editor (β = 1.02, IC95% = 0.31; 1.73, p < 0.001). We found that it takes 11 less days for an editor-in-chief to publish, compared to a non-editor (β = -10.71, IC95% = -8.39; -13.03, p = 0.005).

Distribution of econometric indicators

**Table 2** summarizes the distribution of the three authorship concentration indices. Over the 2020 to 2023 period, the Gini index (*100) median was 12.56 (IQR 8.32 to 18.09), and the 95^th^ percentile was 27.26. Similarly, the Hoover index (*100) median was 12.19 (IQR 8.20 to 17.13), and the 95^th^ percentile was 24.41. The Herfindahl-Hirschman (*10,000) index median was 1.37 (IQR 0.69 to 2.76), and the 95^th^ percentile was 8.41.

## Discussion

It appears that an overrepresentation of the editors of a journal among its authors is far from being exceptional and might be considered a structural property of a whole segment of the biomedical publication field. In this survey of 1,623 journals, with 100,282 editors, we found that in 5% of the cases, a single editor contributes to nearly 7% of a journal’s research articles, excluding editorial publications. The phenomenon isn’t strictly limited to smaller journals. These editors also happen to be editors-in-chief in more than one third of the cases. Nearly half of the most prolific authors have editorial functions within the journal they publish in, and almost 9% are editors-in-chief. Moreover, in half of the journals, 1 out of 8 research article is co-authored by an editor, and **Fig. 1B** shows that this effect is not restricted to smaller journals.

It is also confirmed that certain authors and editors, namely the most prolific, see their manuscripts published faster, compared to regular authors. Publication lags add another dimension to the analysis of disparities between authors with different characteristics, and allows to identify not only differences in the output of the publication process, but also in the form of this process. In 2021, Scanff et al. have shown a significantly shorter publication lag for the most prolific authors compared to the average author, with more than 60% of them being editors (9). We combined two approaches to the publication lag: at journal scale, with the median publication lag, and at article scale, with a linear mixed model. We didn’t find a noticeable difference between authors and editors overall. However, most prolific authors and editors appear to publish faster than their average counterparts, up to 11 days faster in average.

Another aim of the study was to identify situations where disparities didn’t stem from individual authors specifically, but rather from groups of authors. However, it didn’t yield solid conclusions. Our computed Gini indices from journals vary between 0.00 and 0.50, with a median at approximately 0.13. Measuring authorship is obviously different from measuring wealth, and there is no standard about the range within which a Gini index would be considered as indicating a relatively equal treatment between authors. As an illustration, according to the Gini indices available in the World Bank database (19), lowest and highest Gini indices (respectively the Slovak Republic, with 0.24 in 2023, and the Republic of South Africa, with 0.63 in 2014) had a difference of 0.39. This comparison may merely manifest a strong heterogeneity between journals, in their distribution of the volume of authorship among the authors. However, it is difficult to assess what value would be the evidence of a fair distribution and what should be deemed a worrying deviation.

It is of course impossible to *directly* assess the quality and fairness of the evaluation of a set of articles, or of the evaluation practices of a set of journals; yet, quantitative indicators can serve as proxies for *indirectly* identifying deviations from the procedural ideal depicted by the COPE (8), *i.e.* potential conflicts of interest and illegitimate publication biases.

Our dataset shows a clear tendency towards a representation of self-publishing editors amongst the authors in a majority of biomedical journals that goes beyond the spectrum depicted by COPE, and faster publications for the most prolific authors, editors, and particularly editors-in-chief, in a number of journals. This could be the manifestation of different kinds of underlying phenomena, with different consequences. Such disparities could merely reveal a) different levels of competence among the authors: disparities of their overall scientific productivity, literacy of their own field, and acquaintance with the writing conventions of certain journals. An argument for such hypothesis could be that it seems reasonable to expect these three qualities to go together and to give access to editorial responsibilities in the field: experienced researchers would perfectly pre-format their article and anticipate all the common writing mistakes, thus facilitating their reviewers’ task; they would also know better how to position themselves in the field, pursuing or even anticipating research trends that spark the most interest ; none of which prohibiting a precedence to publish in concurrent journal. All this wouldn’t then represent any distortion of the procedural ideal beyond the fact that prioritizing trendy objects could encourage the publication of lower quality yet fashionable studies over dull yet flawless ones.

However, there are strong reasons to also consider a second hypothesis. Such disparities in the share of research articles and publication lags could reveal b) clientelist relations between editors in a significant number of journals. We could define them as the exchange of favors, binding the partners through relations of obligation, on the basis of non-universal rules. For instance, in scientific research, the distribution of resources (access to publication, positions, grants, prestige, etc.) may follow universal rules like the rewarding of merit, of the advancement of scientific knowledge, etc., that is rules that apply to everyone the same way; or it may follow particularistic rules, like prioritizing the family, friends or close colleagues of the one in charge. Our data could then indicate that editors in a number of journals are more inclined to fast-track and to publish their fellow board members – under the presupposition that even if editors registered in January or February 2022, at the moment of the data scraping, have not been during the 2-years interval before, or the 2-year interval after, it is still very plausible that their interpersonal relations with the other editors were existing already or endured afterwards. Reasons supporting such unequal treatments are fairly obvious: it is way more difficult to evaluate someone with whom we have affective bonds or whose success is in our own interest, though we have no way to understand how it is subjectively represented and justified by the self-publishing editors themselves without further qualitative studies.

Hypothesis b) would represent a significant deviation from the ideal publication process, not merely some noise, unavoidable in the effectuation of any ideal form in actual institutions, but a structural bias threatening the integrity of biomedical research. It is true that clientelism is not inherently bad, since it is simply a way to build cohesion in a social formation, yet it contradicts by definition the core norms of science. Practices such as excessive self-citation and self-publication do not by themselves invalidate the results of the studies, but both tend to lean towards publication in support of self (20), are generally associated with a deterioration in the quality of the literature, and are viewed negatively overall (21). Since the advancement of science is far from being only cumulative, the development of breakthrough paradigms sometimes necessitates that smaller communities emerge, granting their members the emulation and mutual support required to transform what could be overlooked as a whimsical idea into a fruitful collaborative endeavor (22). However, the pattern we see emerging in our data isn’t that of a few island communities breaking apart from the main paradigms of biomedical science to lay the foundations of new fields of research, for it is too much distributed across the publishing sphere, even despite strong heterogeneity between journals.

To favor hypothesis b) instead of the very optimistic account of hypothesis a) on empirical grounds rather than on its mere plausibility would require smaller-scale, individually focused studies. Indeed, given the limitations of our current indicators, it remains challenging to provide a detailed account of the social mechanisms underlying this endogamy. Overall, our findings highlight a tension between the idealized publication process depicted by COPE (8), and the social dynamics that embody it, that could threaten the integrity of biomedical research.

## Limitations

Open Editors was the best suited tool for our research interest: having a view of the editorial boards of as much scientific journals as possible. But relying on this dataset highlights a structural problem, namely a single data extraction through web scraping that results in a static dataset. This static data greatly hinders out precision compared to a dynamic dataset. We do not have the capacity to identify potential changes in an editorial board’s composition at a set time. This induces a classification bias, which could be considered non-differential, since only a small proportion of the journals provide the dates of their editors’ mandates. In the absence of centralization – and standardization – of the information of editorial board compositions, we must count on data mining initiatives with clear limitations. Automatized data mining becomes less flexible with an increasing number of journals. This inflexibility comes in several forms, namely poor data standardization due to different norms between the editorial board hierarchy, naming conventions, and difficulties to maintain the tool due to structural code changes in the targeted websites. Our results about editors-in-chief suggest that the information of each editor-in-chief mandate’s beginning and end would be useful to better understand the publication dynamic of a journal, since the average editor-in-chief’s auto-publishing dynamic greatly differs from the average author and editor, even on research articles. Some journals already provide a list of their previous editors-in-chief and their mandate. When taking the (January) 2022 round of scraping from Open Editors, we chose a 2-year interval before and after, with the hypothesis that an editor with effective editorial functions in 2022 would have had an (hypothetical) influence two years before and may keep it two years after. This remains an assumption, as we have no clear way to get the composition of the editorial boards within other time frames. A non-verification of this hypothesis could lead to an overestimation of the editor-related indicators. On the other hand, having only one fixed picture of the editors-in-chief for a time frame of 4 years could lead to an underestimation of the editor-related parameters in general.

The web scraping comes with poor standardization across naming conventions. Thus, we face high risk of mismatching between Open Editors and MEDLINE. These discrepancies led us to establish filters to ensure the association between an editor in OE, and an author in MEDLINE. For an association to be correct, we chose a rather strict limit of a unique mandatory first name initial, and complete last name, between an author and an editor. This approach automatically excluded people who were only referred to in their editorial board with their last name. Besides, we could expect that with these rules, we would be exposed to lower sensibility with countries with a tighter pool of last names. Similarly, the identification of research articles was also set to be restrictive towards contents that matched our definition of research articles, i. e. the research letters were de facto excluded with the inclusion of the Letter format in our negative search string.

The choice of the criteria to define an editor-in-chief was also a key stake, as we chose to exclude associate, deputy and vice editors-in-chief, to get a minimalistic pool of editors-in-chief that evolve clearly at the top of the journal hierarchy. This comes with the consequence that editors-in-chief with lesser responsibilities, as well as associate editors, are considered as regular members of the editorial board within our study. The choice to go only for two classes of editors (editor-in-chief versus non-editor-in-chief) wasn’t motivated by an underlying sociological hypothesis, but by a practical constraint: the diversity of naming conventions amongst the journal roles. This comes with the clear drawback that we can’t verify if there are differences between editors with additional responsibilities who aren’t editors-in-chief, and the regular members of editorial boards.

A final limitation is the relevance of our data in an everchanging scientific community. Our results cease in 2023, due to our limitation to the early 2022 scraping round. Given the critical significance of scientific misconduct for the integrity of the scientific production and the fact that the issue is gaining momentum in the academic and public sphere, the introduction of public policies aiming at preventing it or mitigating its effects may very well have altered publication patterns and hindered nepotistic behaviors.

Our results will be complemented by a qualitative approach to examine whether research articles authored by editors appropriately address potential conflicts of interest, as required by COPE recommendations, and to better understand the motivations underlying this practice. This issue was already studied for authors of Cochrane reviews, when authors were involved in the editorial board. An appropriate declaration of the conflict was reported in less than 5% of the cases (23).

## Data Availability

Data, codes and materials supporting the study will be uploaded to the Open Science Framework: https://doi.org/10.17605/OSF.IO/EYXH7.

## Appendix 1: Supplementary figures

**Supplementary figure 1:**
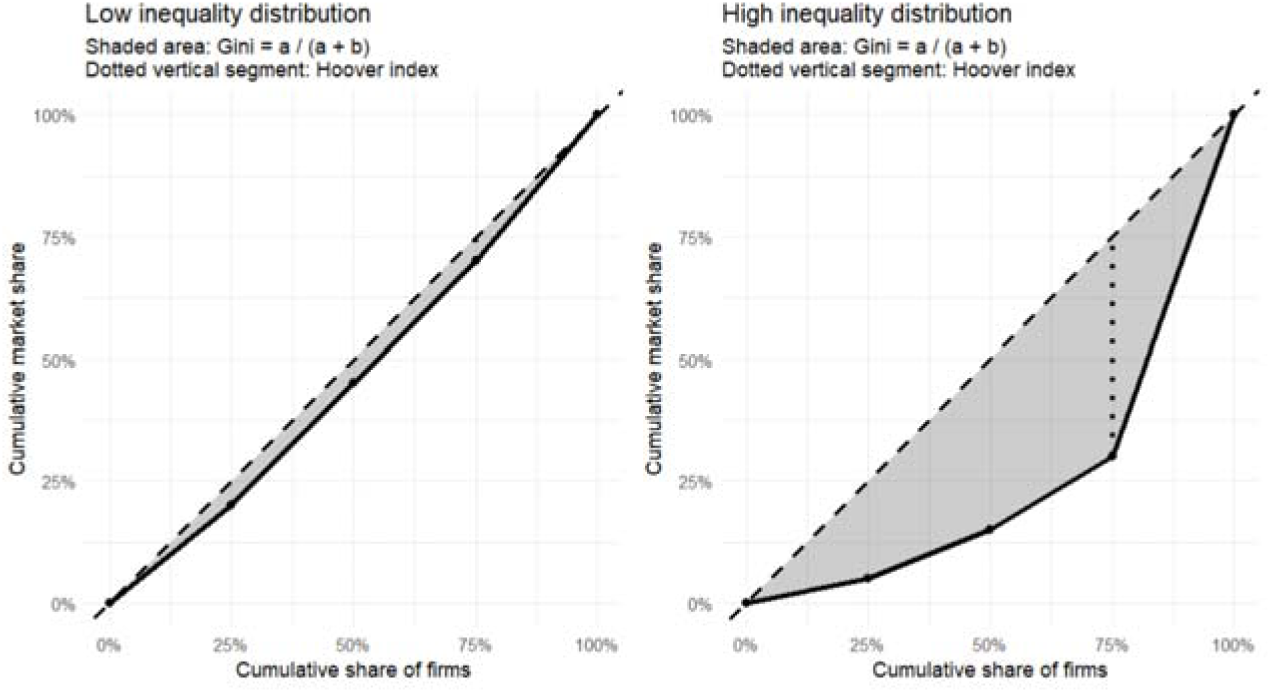
Graphical representations of the Gini index.

**Supplementary figure 2:**
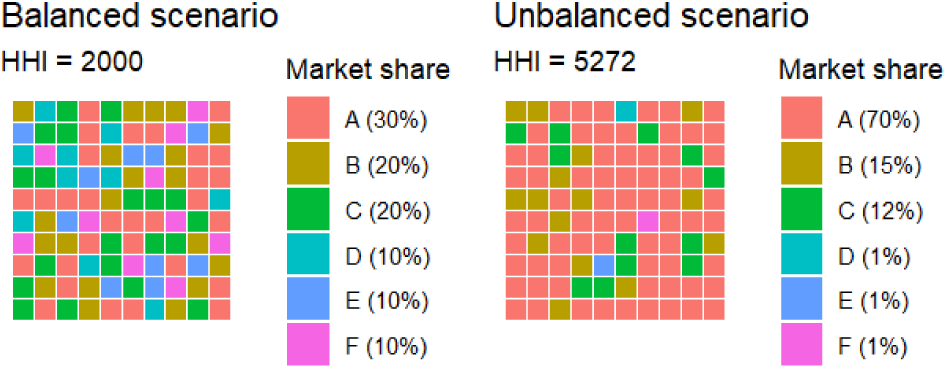
Graphical representations of the Herfindahl-Hirschman index.

**Supplementary figure 3:**
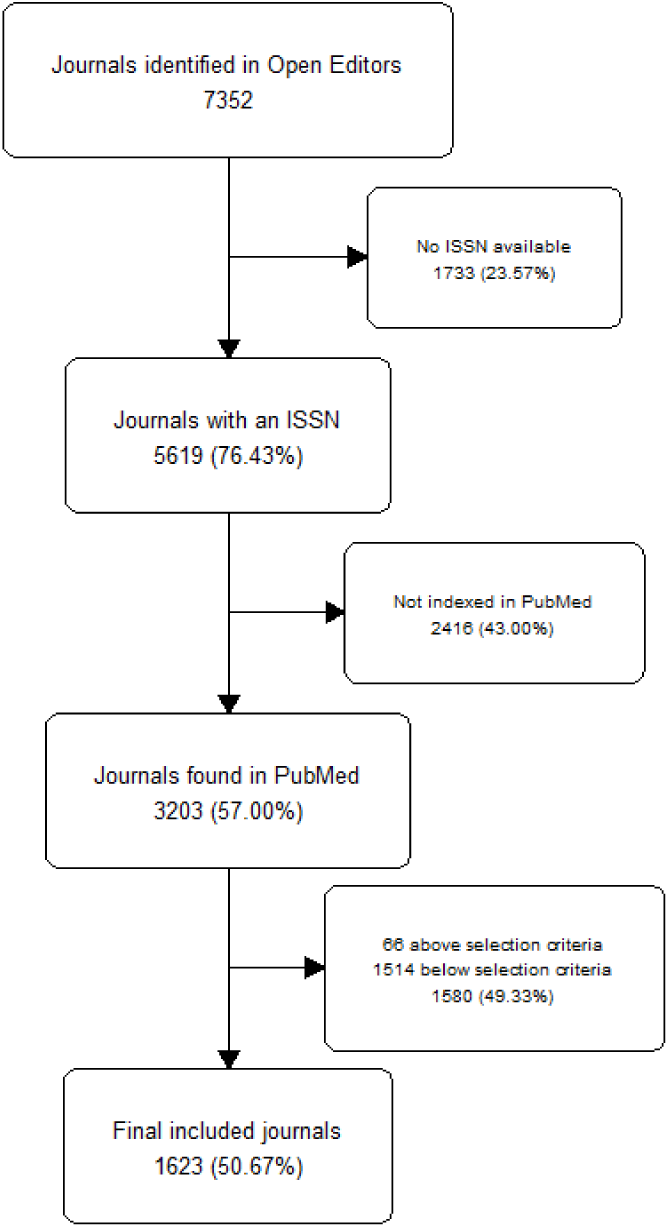
Flow chart of journal selection.

## APPENDIX 2: STROBE Statement—checklist of items that should be included in reports of observational studies

abc: not applicable; abc: recommendation was not followed; abc: recommendation was followed

|  | Item No | Recommendation |
| --- | --- | --- |
| **Title and abstract** | 1 | (a) Indicate the study's design with a commonly used term in the title or the abstract<br>(b) Provide in the abstract an informative and balanced summary of what was done and what was found |
| **Introduction** |  |  |
| Background/rationale | 2 | Explain the scientific background and rationale for the investigation being reported |
| Objectives | 3 | State specific objectives, including any prespecified hypotheses |
| **Methods** |  |  |
| Study design | 4 | Present key elements of study design early in the paper |
| Setting | 5 | Describe the setting, locations, and relevant dates, including periods of recruitment, exposure, follow-up, and data collection |
| Participants | 6 | (a) *Cohort study*—Give the eligibility criteria, and the sources and methods of selection of participants. Describe methods of follow-up<br>*Case-control study*—Give the eligibility criteria, and the sources and methods of case ascertainment and control selection. Give the rationale for the choice of cases and controls<br>*Cross-sectional study*—Give the eligibility criteria, and the sources and methods of selection of participants<br>(b) *Cohort study*—For matched studies, give matching criteria and number of exposed and unexposed<br>*Case-control study*—For matched studies, give matching criteria and the number of controls per case |
| Variables | 7 | Clearly define all outcomes, exposures, predictors, potential confounders, and effect modifiers. Give diagnostic criteria, if applicable |
| Data sources/measurement | 8* | For each variable of interest, give sources of data and details of methods of assessment (measurement). Describe comparability of assessment methods if there is more than one group |
| Bias | 9 | Describe any efforts to address potential sources of bias |
| Study size | 10 | Explain how the study size was arrived at |
| Quantitative variables | 11 | Explain how quantitative variables were handled in the analyses. If applicable, describe which groupings were chosen and why |
| Statistical methods | 12 | (a) Describe all statistical methods, including those used to control for confounding<br>(b) Describe any methods used to examine subgroups and interactions<br>(c) Explain how missing data were addressed<br>(d) *Cohort study*—If applicable, explain how loss to follow-up was addressed<br>*Case-control study*—If applicable, explain how matching of cases and controls was addressed<br>*Cross-sectional study*—If applicable, describe analytical methods taking account of sampling strategy<br>(e) Describe any sensitivity analyses |
| **Results** |  |  |
| Participants | 13* | (a) Report numbers of individuals at each stage of study—eg numbers potentially eligible, examined for eligibility, confirmed eligible, included in the study, completing follow-up, and analysed<br>(b) Give reasons for non-participation at each stage<br>(c) Consider use of a flow diagram * in supplementary |
| Descriptive data | 14* | (a) Give characteristics of study participants (eg demographic, clinical, social) and information on exposures and potential confounders<br>(b) Indicate number of participants with missing data for each variable of interest<br>(c) *Cohort study*—Summarise follow-up time (eg, average and total amount) |
| Outcome data | 15* | *Cohort study*—Report numbers of outcome events or summary measures over time<br>*Case-control study*—Report numbers in each exposure category, or summary measures of exposure<br>*Cross-sectional study*—Report numbers of outcome events or summary measures |
| Main results | 16 | (a) Give unadjusted estimates and, if applicable, confounder-adjusted estimates and their precision (eg, 95% confidence interval). Make clear which confounders were adjusted for and why they were included<br>(b) Report category boundaries when continuous variables were categorized<br>(c) If relevant, consider translating estimates of relative risk into absolute risk for a meaningful time period |
| Other analyses | 17 | Report other analyses done—eg analyses of subgroups and interactions, and sensitivity analyses |
| **Discussion** |  |  |
| Key results | 18 | Summarise key results with reference to study objectives |
| Limitations | 19 | Discuss limitations of the study, taking into account sources of potential bias or imprecision. Discuss both direction and magnitude of any potential bias |
| Interpretation | 20 | Give a cautious overall interpretation of results considering objectives, limitations, multiplicity of analyses, results from similar studies, and other relevant evidence |
| Generalisability | 21 | Discuss the generalisability (external validity) of the study results |
| **Other information** |  |  |
| Funding | 22 | Give the source of funding and the role of the funders for the present study and, if applicable, for the original study on which the present article is based |

\*Give information separately for cases and controls in case-control studies and, if applicable, for exposed and unexposed groups in cohort and cross-sectional studies.
**Note:** An Explanation and Elaboration article discusses each checklist item and gives methodological background and published examples of transparent reporting. The STROBE checklist is best used in conjunction with this article (freely available on the Web sites of PLoS Medicine at http://www.plosmedicine.org/, Annals of Internal Medicine at http://www.annals.org/, and Epidemiology at http://www.epidem.com/). Information on the STROBE Initiative is available at www.strobe-statement.org.

## APPENDIX 3: Filters

APPENDIX 3: Filters A. Research articles
|  | Outcome + | Outcome - |
| --- | --- | --- |
| Test + | 87 | 0 |
| Test - | 2 | 11 |

B. Disambiguation
|  | Outcome + | Outcome - |
| --- | --- | --- |
| Test + | 106 | 0 |
| Test - | 3 | 11 |

C. Editors-in-chief
|  | Outcome + | Outcome - |
| --- | --- | --- |
| Test + | 105 | 2 |
| Test - | 8 | 391 |

Sensitivity: 0,93 (0,87; 0,97)
**Specificity:** 0,99 (0,98; 1,00)
Predictive positive value: 0,99 (0,93; 1,00)
Predictive negative value: 0,98 (0,96; 0,99)

